# Alzheimer’s disease-related cerebrospinal fluid Biomarkers in Amyotrophic Lateral Sclerosis: Real-World Associations with Disease Status, Cognition and Tofersen Response

**DOI:** 10.64898/2026.10.26.26366525

**Authors:** Felicitas Eberle, Sarah Bernsen, Rachel Fabian, Catherine Widmann, Patrick Weydt, Sergio Castro-Gomez

**Author notes:** Correspondence to: Sergio Castro-Gomez MD, PhD or Patrick Wydt MD Center for Neurology, Venusberg-Campus 1, University Hospital Bonn, University of Bonn, 53129 Bonn, Germany. These authors contributed equally to this work (Felicitas Eberle, Sergio Castro-Gomez).

## Abstract

**Background and Objectives:** Amyotrophic lateral sclerosis (ALS) is a neurodegenerative disease primarily affecting motor neurons but also frequently involving cognitive impairment. Cerebrospinal fluid (CSF) measurements of β-amyloid peptides (Aβ_40_, Aβ_42_), tau proteins, and neurofilaments are clinically established biomarkers in Alzheimer’s disease (AD). Emerging evidence suggests that AD-related pathological changes and AD-related biomarkers may appear in ALS, yet their diagnostic and prognostic significance remains incompletely understood.

**Methods:** In this retrospective study, we analyzed clinical and CSF biomarker data from patients with ALS, AD, Huntington’s disease (HD), and other disease controls from real-world clinical settings. The dataset comprised Aβ_40_, Aβ_42_, total tau (t-Tau), phosphorylated tau181 (pTau181), phosphorylated neurofilament heavy chain (pNfH), and relevant serum markers as well as clinical and cognitive status measurements. Additionally, longitudinal CSF sampling over one year was evaluated in three ALS patients receiving tofersen treatment.

**Results:** In this real-world cohort, AD-related CSF biomarkers demonstrated no diagnostic value for distinguishing ALS from other neurodegenerative conditions. However, Aβ_40_, Aβ_42_, and pTau181 levels correlated positively with ALS Functional Rating Scale (ALSFRS-R) global score and serum Creatine Kinase MB (CK-MB) concentrations. At the same time, a higher Aβ_42_/Aβ_40_ ratio was associated with better memory performance on the Edinburgh Cognitive and Behavioural ALS Screen (ECAS). In tofersen-treated cases, AD-related biomarkers gradually declined during follow-up.

**Discussion:** These findings suggest that elevated CSF AD–associated biomarkers in ALS may reflect preserved functional and cognitive status and exhibited parallel declines during follow-up in three tofersen-treated patients, supporting their further prospective evaluation as exploratory markers of therapeutic responsiveness.

**Graphical Abstract:** 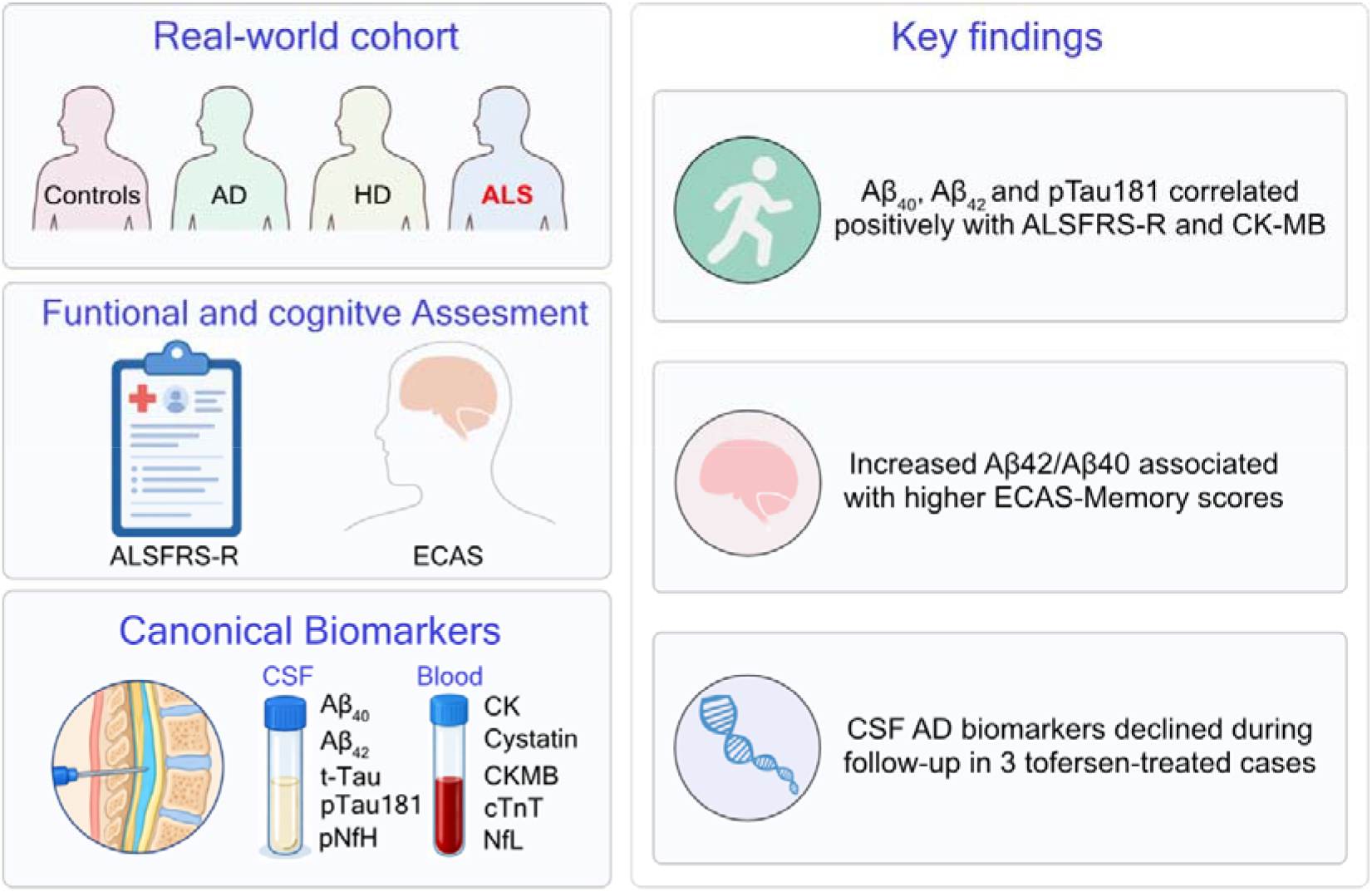

## Introduction

Amyotrophic lateral sclerosis (ALS) manifests as a rapid, progressive, fatal neurodegeneration marked by degeneration of motor neurons within the motor cortex, brainstem, and spinal cord [1, 2]. Despite major advances in understanding its molecular and genetic mechanisms, early and reliable diagnosis remains challenging. This is largely due to clinical heterogeneity of ALS and the absence of specific biomarkers that can distinguish ALS from other neurodegenerative conditions, guide therapeutic decision-making, and facilitate patient stratification and follow-up in clinical trials [1]. Currently, diagnosis of ALS primarily depends on clinical evaluation aligned with established criteria and complemented by electromyography to confirm lower motor neuron involvement [3–5]. To characterize disease severity and cognitive involvement, standardized instruments including the ALS Functional Rating Scale–Revised (ALSFRS-R) [6] and the Edinburgh Cognitive and Behavioral ALS Screen (ECAS) are widely applied [7]. The measurement of Neurofilament light chain (NfL) [8, 9] and phosphorylated neurofilament heavy chain (pNfH) [10, 11] in blood and cerebrospinal fluid (CSF) serve as indicators of neuroaxonal damage, with levels substantially raised in ALS relative to healthy individuals. However, neurofilament elevations lack specificity for ALS, appearing likewise elevated in other neurological disorders such as frontotemporal dementia (FTD), multiple sclerosis, Creutzfeldt-Jakob disease, and Alzheimer’s disease (AD) [12]. Regarding the latter, evidence from imaging and postmortem analyses reveals concomitant amyloid-β and tau aggregates in ALS neural tissue, hinting at overlapping Alzheimer-type pathologies [13, 14]. Investigations of CSF amyloid-β isoforms (Aβ_40_, Aβ_42_), total tau (t-Tau), and phosphorylated tau at serine 181 (pTau181) in ALS remain incomplete, though these may also nsignal synaptic loss and axonal harm [15]. Moreover, a recent report using target proteomics suggested that CSF AD-related biomarkers may respond to Disease-Modifying Therapies (DMT) such as tofersen [16]. Tofersen, an antisense oligonucleotide delivered intrathecally, targets Superoxide Dismutase 1 (*SOD1*) mRNA to curb protein production in ALS patients with *SOD1* mutations, yielding reductions in CSF SOD1 and neurofilaments in trial cohorts [17]. This intervention offers a lens to assess if SOD1 lowering alters trajectories of CSF and blood biomarkers, and it is the first model for DMT in ALS.

In this study, we conducted a systematic analysis of CSF concentrations of Aβ_40_, Aβ_42_, t-Tau, and pTau_181_ in patients with ALS and compared them with concentrations in disease controls and patients with other neurodegenerative disorders, including AD and Huntington’s disease (HD), in a real-world clinical setting. Our first objective was to evaluate the diagnostic discriminatory capacity of these biomarkers across the disease groups. Our second objective was to determine, within the ALS collective, whether CSF biomarker concentrations were associated with clinical functional status, muscle-derived markers including creatine kinase myocardial band (CK-MB), neurofilament concentrations, and cognitive performance. Finally, as an exploratory analysis, we assessed longitudinal changes in these biomarkers during tofersen treatment in patients with genetically confirmed *SOD1*-associated ALS and examined their relationship with neurofilament dynamics.

## Materials and methods

### Study participants

Clinical data were retrospectively retrieved from the electronic medical records of the Center for Neurology and the former Clinic for Neurodegenerative Diseases at University Hospital Bonn, Germany. The study included individuals diagnosed with Alzheimer’s disease (AD), Amyotrophic Lateral Sclerosis (ALS) or Huntington’s disease (HD), as well as individuals with conditions considered clinically non-neurodegenerative, including depression, polyneuropathy, lumbar pain and normal pressure hydrocephalus. We refer to the latter group as non-neurodegenerative disease (non-NND) controls to distinguish them from neurodegenerative disease (NND) groups. All participants underwent neurological evaluations between October 2019 and August 2021. Data were obtained from routine diagnostic and follow-up assessments; no additional procedures or interventions were performed for this study.

A total of 52 individuals met the Revised El Escorial criteria for ALS, based on comprehensive clinical evaluation, medical history, and supporting diagnostic investigations [18]. None of the patients with ALS in this collective fulfilled the clinical criteria for frontotemporal dementia (FTD). The other NND groups comprised 17 patients with genetically confirmed HD, defined by an abnormally expanded CAG repeat in the *HTT* gene (>39 repeats), and 55 patients with AD, characterized by typical clinical features and a CSF biomarker profile consistent with AD (amnestic mild cognitive impairment or dementia, a reduced Aβ_42_/Aβ_40_ ratio and increased pTau181). In addition, 48 individuals with clinically non-neurodegenerative conditions were included as disease controls, as previously described. The date of lumbar puncture was used as the reference time point for data collection and clinical and functional assessment. Measurements were included only if obtained within a prespecified time window (±15 days) around this time point. Accordingly, data were not available for all participants for selected assessments and biomarkers: ALSFRS-R scores were available for 40 patients (76.9%), pNfH measurements for all 52 patients (100%), CK-MB levels for 48 patients (92.3%), and ECAS-Memory Score assessments for 33 patients (63.5%) (Supplementary Table 1. Availability Table).

Among the patients with ALS, three had a confirmed *SOD1* mutation and received tofersen treatment between January 2025 and January 2026. These patients continued to attend regular follow-up visits. Laboratory measurements were part of routine clinical care and were analyzed retrospectively. The study was approved by the Ethics Review Board of the University of Bonn, Germany (file no. 2026-540-BO).

### Data collection and samples measurements

The data used in this study comprised clinical information and biomarker values obtained from biological samples (CSF and serum) collected as part of routine medical care. The general clinical dataset for ALS patients included gender, diagnosis, age at onset, first clinical presentation, and the site and time of initial manifestation. ALSFRS-R[19] and ECAS[20] assessments were performed within a few days before or after lumbar puncture (maximum ±15 days). In addition, standard laboratory biomarkers routinely analyzed in our routine practice and initial evaluation, such as pNfH in CSF, creatinine, reactive protein C (RPC), cystatin C, creatine kinase (CK), CK-MB, N-terminal prohormone of brain natriuretic peptide (NT-proBNP), and cardiac troponin T (cTnT)— were included. While the above was primarily carried out for ALS patients, the results for AD-related biomarkers (Aβ_40_, Aβ_42_, t-Tau, pTau181 as well as the Aβ_42_/Aβ_40_ and the Aβ_42_/p-Tau ratios) were compiled for the entire patient cohort. A pseudonymized and anonymized database was created from the collected data to perform statistical analysis in compliance with the current applicable European and German data protection laws.

Routine serum and plasma chemistry was performed in the certified central diagnostic laboratory at the Institute of Clinical Chemistry and Clinical Pharmacology, University Hospital Bonn, Germany, using commercially available assay kits. Quantification of pNfH in CSF was carried out at the University Medical Center Ulm, Germany, with the ELLA microfluidic system (Bio-Techne, Minneapolis, USA)[21]. Measurement of AD-related biomarkers was performed using the LUMIPULSE G600II automated platform (Fujirebio, Japan) at the German Centre for Neurodegenerative Diseases as part of the routine paraclinical evaluation[22].

### Statistical analysis

Statistical analyses were performed using GraphPad Prism version 9.5 (GraphPad Software, San Diego, CA, USA), and figures were prepared using Affinity by Canva (Sydney, Australia). Distributions were assessed using the Shapiro–Wilk test and quantile–quantile (Q–Q) plots. Because most variables were not normally distributed, unadjusted between-group comparisons were performed using the Kruskal–Walli’s test, followed by Dunn’s multiple comparisons test. Associations between AD-related biomarkers and ALS-related laboratories and clinical measures were initially evaluated using Spearman’s rank correlation. Specifically, Aβ_40_, Aβ_42_, t-Tau, pTau_181_, the Aβ_42_/Aβ_40_ ratio, and the Aβ_42_/p-Tau ratio were correlated with relevant laboratory parameters and clinical progression measures, including the ALSFRS-R and ECAS.

In the three patients with SOD1-associated ALS receiving tofersen, longitudinal CSF concentrations of AD-related biomarkers (Aβ_40_, Aβ_42_, t-Tau, pTau181) and neurofilaments were examined descriptively. Biomarker values were naturally log-transformed for visualization, and individual trajectories were plotted. First and last available follow-up values were summarized for each patient. Given the small sample size, analyses focused on the direction and magnitude of within-patient changes rather than formal hypothesis testing. These exploratory observations are not interpreted as evidence of a treatment effect and were intended to generate hypotheses for future prospective studies in larger cohorts. All tests were two-sided, with a nominal significance threshold of *p* < 0.05. Effect estimates are reported with 95% confidence intervals (CIs), where applicable.

## Results

### Cohort characteristics and Biomarker Comparisons

In this retrospective study, we reviewed medical records from the former Clinic of Neurodegenerative Diseases and the Centre for Neurology at University Hospital Bonn, Germany. The cohort comprised 172 patients with neurodegenerative or non-neurodegenerative disorders (51% female), classified into four diagnostic groups: non-neurodegenerative controls (non-NND, Control; *n* = 48), Alzheimer’s disease (AD; *n* = 55), Huntington’s disease (HD; *n* = 17), and amyotrophic lateral sclerosis (ALS; *n* = 52). Demographic characteristics and biomarker concentrations are summarized in Table 1 as medians (ranges). Aβ_40_ concentrations were normally distributed across groups; all other variables showed lognormal or non-normal distributions. The AD group was significantly older than the other groups (P < 0.0001) and included a higher proportion of women, whereas patients with HD were younger than controls (*p* = 0.0008).

**Table 1.** AD biomarker in the real-world cohort—University Hospital Bonn.

|  |  |  |  |  |  | **ALS vs Control** |  |
|---|---|---|---|---|---|---|---|
|  | **Control (n = 48)** | **AD (n = 55)** | **HD (n = 17)** | **ALS (n = 52)** | ***P*-value** | **Effect r** | ***P* post hoc** |
| Age | 66<br>(27 - 80) | 81<br>(45 - 95) | 54<br>(29 - 77) | 67<br>(44 – 84) | **<0.0001<sup>a</sup>** | 0.64 | >0.9999 |
| Sex (f:m) | 20:28 | 39:16 | 8:9 | 21:31 | **0.0050<sup>b</sup>** | Cramers V = 0.27 | 0.7400 |
| Aβ<sub>40</sub> (pg/ml) | 13714.00 (4480.00 - 21023.00) | 13453.00<br>(6315.00 - 23428.00) | 14151.00<br>(7470.00 - 22876.00) | 12.148<br>(4358.00 - 22337.00) | **0.4277<sup>c</sup>** | 0.14 | >0.9999 |
| Aβ<sub>42</sub> (pg/ml) | 1066.00<br>(396.00 – 2208.00) | 598.00<br>(231.00 - 2049.00) | 1487.00<br>(712.00 - 2335.00) | 1150.00<br>(439.00 - 2335.00) | **<0.0001<sup>a</sup>** | 0.66 | > 0.9999 |
| t-Tau (pg/ml) | 235.50<br>(74.00 – 818.00) | 651.00<br>(228.00 – 1589.00) | 291.00<br>(142.00 – 441.00) | 239.00<br>(94.00 – 866.00) | **<0.0001<sup>a</sup>** | 0.66 | > 0.9999 |
| pTau181 (pg/ml) | 39.25<br>(15.80 - 144.30) | 102.60<br>(34.00 - 283.10) | 39.70<br>(21.80 - 75.80) | 33.95<br>(12.40 - 145.10) | **<0.0001<sup>a</sup>** | 0.71 | > 0.9999 |
| Aβ<sub>42</sub>/Aβ<sub>40</sub> | 0.10<br>(0.03 - 0.12) | 0.05<br>(0.02 - 0.11) | 0.10<br>(0.09 - 0.11) | 0.10<br>(0.03 - 0.12) | **<0.0001<sup>a</sup>** | 0.72 | 0.5263 |
| Aβ<sub>42</sub>/pTau181 | 35.19<br>(4.01 - 51.37) | 5.50<br>(1.86 - 43.05) | 38.58<br>(26.5 - 52.15) | 35.50<br>(4.71 - 57.8) | **<0.0001<sup>a</sup>** | 0.75 | > 0.9999 |

<sup>a</sup>Kruskal-Wallis Test
<sup>b</sup>Chi-square
<sup>c</sup>One-Way ANOVA
Aβ, Amyloid β; AD, Alzheimer`s Disease; ALS; Amyotrophic Lateral Sclerosis; HD, Huntington’s Disease; t-Tau, total Tau, pTau<sub>181</sub>, phosphorylated Tau in Serine 181

ALS patients were stratified by site of onset into bulbar-onset (n = 13) and spinal-onset (n = 39) subgroups. Clinical, biomarker, and cognitive data are presented as medians (ranges) in Table 2. As expected, the bulbar-onset subgroup had a significantly lower body mass index (P = 0.025) and ALS Functional Rating Scale– Revised (ALSFRS-R) total score (P = 0.013), consistent with greater clinical impairment (Table 2). Patients with concomitant frontotemporal dementia (ALS-FTD) were not represented in this cohort.

**Table 2.** Characterization of ALS patient real-world cohort.

|  |  | **ALS subgroups** |  |  |
|---|---|---|---|---|
|  | **ALS Total (n = 52)** | **Bulbar onset (n = 13)** | **Spinal onset (n = 39)** | ***P*-value** |
| **Sex, (female: male)** | 21:31 | 5:8 | 16:23 | >0.999 |
| **Age, years** | 67 (42 - 84) | 68.50 (59.00 – 82.00) | 63.00 (42.00 – 84.00) | 0.1639 |
| **BMI (kg/m<sup>2</sup>)** | 24.01 (15.60 - 62.50) | 21.60 (17.36 - 26.72) | 25.74 (15.60 - 62.50) | **0.025<sup>a</sup>** |
| **Duration, years** | 6.00 (0.00 – 16.00) | 5.50 (1.00 – 11.00) | 6.00 (0.00 – 16.00) | 0.306 |
| **Aβ<sub>40</sub> (pg/ml)** | 12.148<br>(4358.00 - 22337.00) | 10480 (7406 - 21317) | 12148 (4358 - 22337) | 0.5881 |
| **Aβ<sub>42</sub> (pg/ml)** | 1150.00<br>(439.00 - 2335.00) | 1006 (509 - 2258) | 1181 (421 - 2335) | 0.7026 |
| **t-Tau (pg/ml)** | 239.00<br>(94.00 – 866.00) | 271.00 (174.00 – 534.00) | 232.50 (94.00 – 866.00) | 0.3538 |
| **pTau181 (pg/ml)** | 33.95<br>(12.40 - 145.10) | 30.30 (22.20 – 99.50) | 34.70 (12.40 – 145.10) | 0.7553 |
| **Aβ<sub>42</sub>/Aβ<sub>40</sub>** | 0.10<br>(0.03 - 0.12) | 0.10 (0.04 – 0.10) | 0.10 (0.03 – 0.12) | 0.8462 |
| **Aβ<sub>42</sub>/pTau181** | 35.50<br>(4.71 - 57.80) | 36.04 (5.12 – 46.18) | 34.97 (4.71 – 57.80) | 0.9430 |
| **CSF pNfH (pg/ml)** | 2263.00 (188 - 11327) | 3362(702 - 11327) | 2015 (241 - 10667) | 0.067 |
| **NfL (pg/ml)** | 132.00 (21 - 699) | 152.00 (54 - 699) | 91.00 (21 - 224) | 0.168 |
| **Cystatin C** | 0.96 (0.62 - 1.55) | 1.07 (0.62 - 1.38) | 0.94 (0.74 - 1.55) | 0.636 |
| **CK (U/L)** | 152.00 (34 - 1848) | 128.00 (43 - 605) | 161.5 (34 - 1848) | 0.349 |
| **CK-MB (ng/ml)** | 5.65 (1.00 -19.20) | 6.20 (1.00 - 19.20) | 5.50 (1.30 - 18.60) | 0.868 |
| **cTnT (ng/l)** | 15.70 (3.70 - 98.30) | 15.70 (4.60 - 66.20) | 15.55 (3.70 - 98.30) | 0.761 |
| **D50** | 16.00 (0.39 - 81.60) | 16.80 (8.00 - 34.00) | 15.50 (0.39 - 81.60) | 0.895 |
| **ALSFRS-R global** | 37.00 (0 - 48) | 31.50 (21 - 41) | 38.50 (10 - 48) | **0.013<sup>a</sup>** |
| **ALSFRS-R bulbar** | 11.00 (2 - 12) | 7.50 (2 - 12) | 12.00 (3 - 12) | **0.002<sup>a</sup>** |
| **ALSFRS-R motorik** | 9.00 (0 - 12) | 6.5 (3 - 12) | 9.00 (0 - 12) | 0.239 |
| **ALSFRS-R mobility** | 8.00 (0-12) | 6.00 (4 - 12) | 8.00(0 - 12) | 0.227 |
| **ALSFRS-R breathing** | 12.00 (2 - 12) | 10.00 (7 - 12) | 12.00 (2 - 12) | **0.025<sup>a</sup>** |
| **ECAS Global** | 108.00 (34 - 125) | 102.00 (34 -110) | 108.00 (45 - 125) | 0.154 |
| **ECAS ALS-S** | 79.00 (24 - 93) | 76.00 (24 -84) | 80.00 (34 -93) | 0.199 |
| **ECAS ALS-NS** | 28.00 (10 - 33) | 25.00 (10 - 31) | 28.00 (11 -33) | 0.132 |
| **ECAS Language** | 26.00 (14 - 28) | 26.00 (14 – 27) | 26.00 (15 – 28) | 0.288 |
| **ECAS Speech** | 20.00 (0 - 24) | 19.00 (4 - 20) | 20.00 (0 - 24) | 0.564 |
| **ECAS Executive** | 35.00 (10 - 42) | 30.00 (10 - 37) | 35.50 (14 - 42) | 0.138 |
| **ECAS Memory** | 16.50 (1 - 21) | 14.00 (1 -19) | 17.00 (4 - 21) | 0.169 |
| **ECAS Spatial** | 12.00 (4 -12) | 11.00 (9 - 12) | 12.00 (4 - 12) | 0.075 |

<sup>a</sup>Mann-Whitney test
Aβ, Amyloid β; ALS; Amyotrophic Lateral Sclerosis; ALSFRS-R, ALS Functional Rating Scale–Revised; BMI, Body Mass Index; ECAS, Edinburgh Cognitive and Behavioral ALS Screen; ECAS ALS-S, ECAS ALS-Specific, ECAS ALS-NS, ECAS ALS non-specific; t-Tau, total Tau, pTau181, phosphorylated Tau in Serine 181;

CSF AD biomarkers (Aβ_40_, Aβ_42_, t-Tau, pTau181, Aβ_42_/Aβ_40_ ratio, Aβ_42_/pTau181 ratio) were compared across diagnostic groups using Kruskal–Wallis tests. As expected, compared with non-NND, patients with AD had significantly lower Aβ_42_ (P = 0.0062, Fig), higher pTau181 (*p* < 0.0001), and lower Aβ_42_/Aβ_40_ and Aβ_42_/pTau181 ratios (both *p* < 0.0001). By contrast, biomarker levels in the ALS and HD groups were comparable to those in controls. These findings support the specificity of CSF AD biomarkers for distinguishing AD from the other diagnostic groups and do not suggest diagnostic value for ALS or HD.

**Figure 1.**
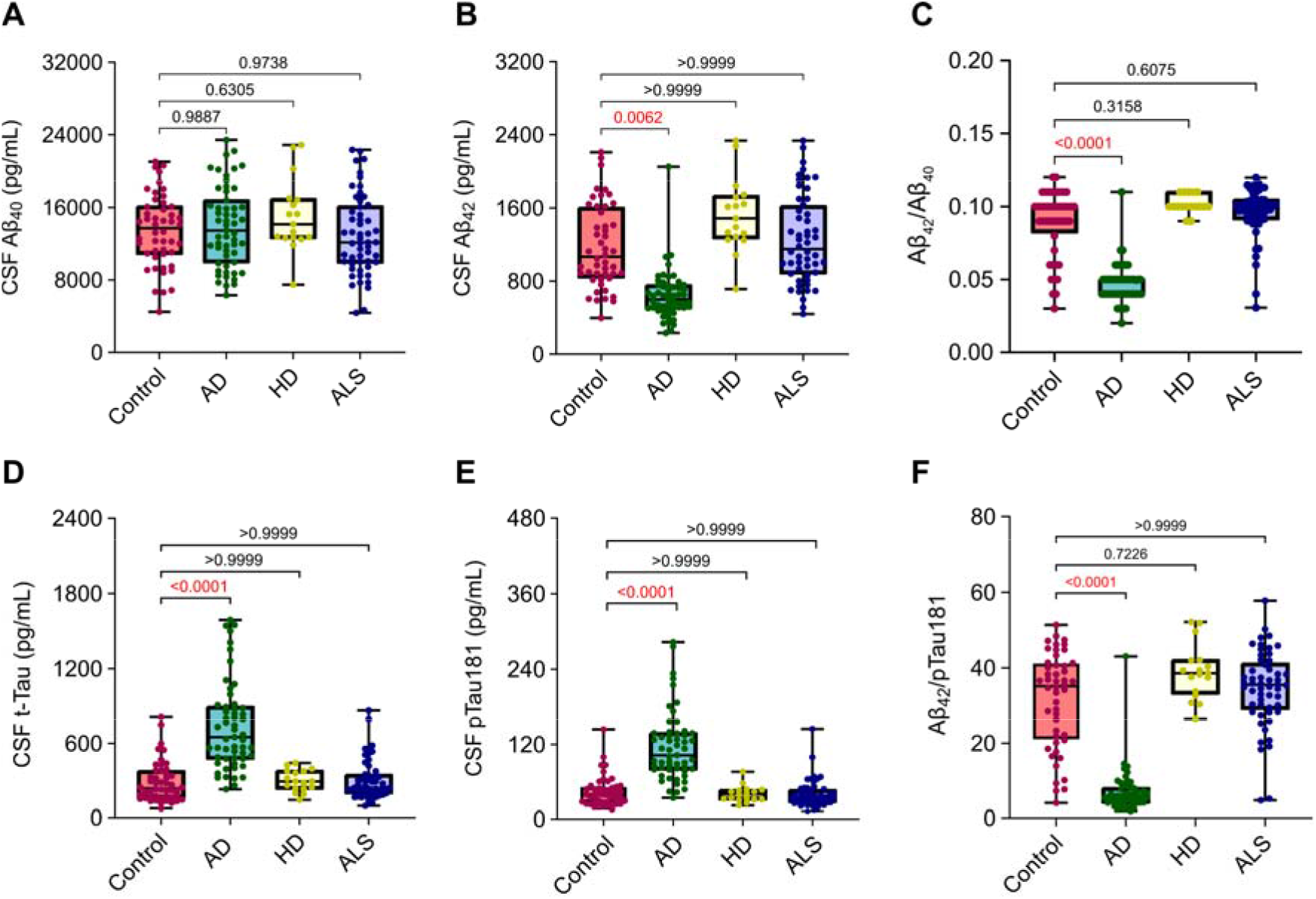
CSF AD Biomarkers levers in ALS and other disorders. CSF levels of (**A**) Aβ_40_, (**B**) Aβ_42_, (**C**) Aβ_42_/Aβ_40_ ratio, (**D**) t-Tau, (**E**) pTau181, and (**F**) Aβ_42_/pTau181 ratio in ALS, non-NND, and NDD (AD and HF) groups. Each dot represents an individual patient. Box plots show median values, with whiskers indicating minimum and maximum values. Reported *P* is from Kruskal–Wallis tests.

### CSF AD biomarkers correlate with ALSFRS-R global score and Muscle Markers

To assess whether CSF AD biomarkers were associated with disease severity or functional impairment in ALS, we performed Spearman correlation analyses with ALSFRS-R (Figure 2A–F) scores and other muscle markers such as CK-MB levels (Figure 3A-F). For ALSFRS-R, we included the assessment closest to CSF collection when available (n = 40; Figure 2A–F); the number of observations for CK-MB varied according to availability (n = 48; Figure 3A–F). Higher ALSFRS-R global scores correlated positively with Aβ_40_ (ρ = 0.391, P = 0.01578; Figure 2A), Aβ_42_ (ρ = 0.369, P = 0.02833; Figure 2B), and mildly with pTau181 (ρ = 0.319, P = 0.03960; Figure 2E). CK-MB levels correlated positively with Aβ_40_ (ρ = 0.438, P = 0.001636; Figure 3A), Aβ_42_ (ρ = 0.478, P = 0.0006; Figure 3B), t-Tau (ρ = 0.297, P = 0.0405,; Figure 3D), and pTau181 (ρ = 0.355, P = 0.0134; Figure 3E). No significant associations were observed between CK-MB and the biomarker ratios (Figure 2A–F). These findings suggest that CSF AD-related proteins are associated with measures of functional status and peripheral disease involvement in ALS.

**Figure 2.**
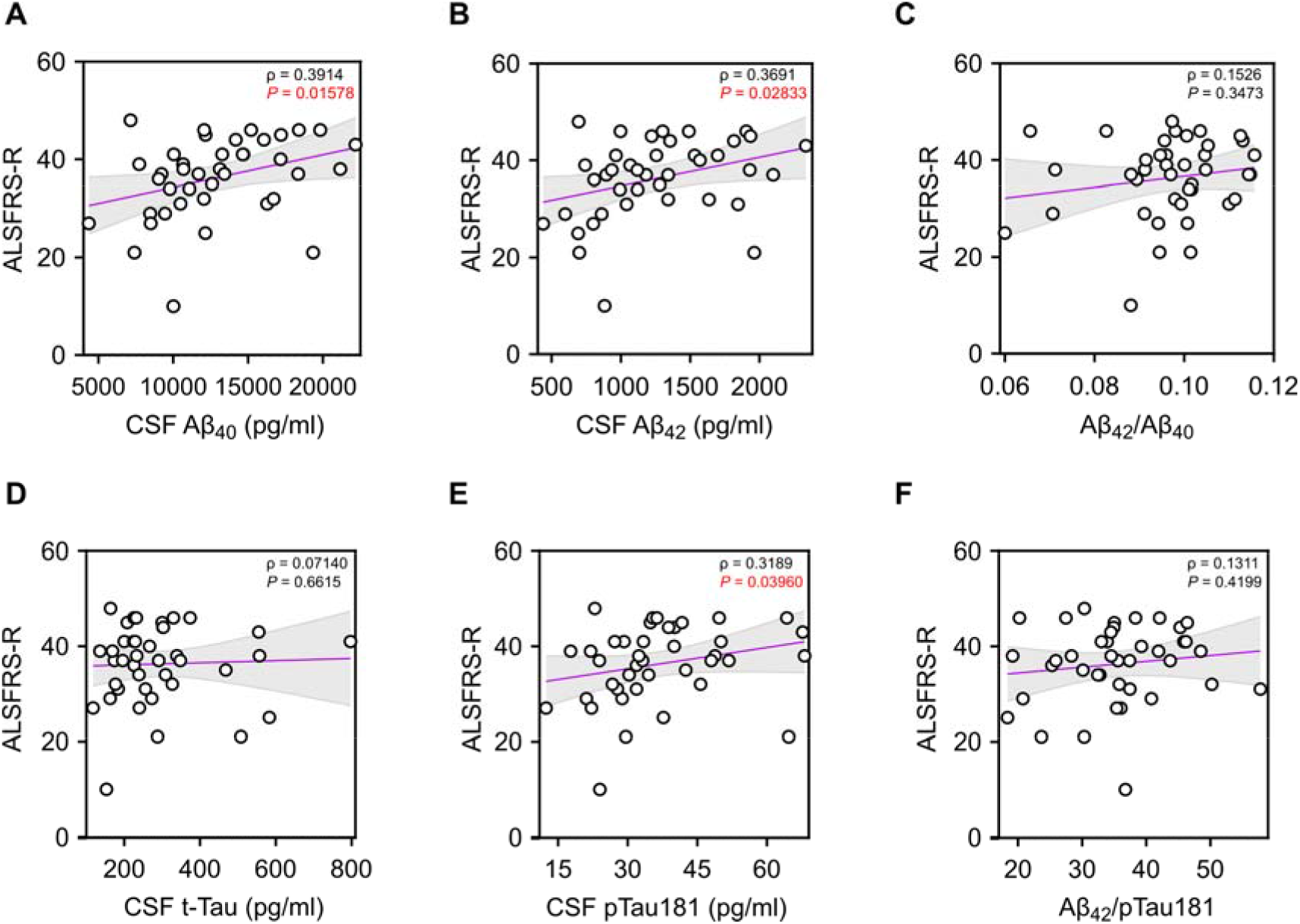
CSF AD Biomarkers Correlations with ALS Functional Scores. Correlation analyses were performed using nonparametric Spearman correlations (ρ). Curves were drawn by a linear regression model with an interaction term for ALSFRS-R Global Scores by CSF levels of (**A**) Aβ_40_, (**B**) Aβ_42_, (**C**) Aβ_42_/Aβ_40_ratio, (**D**) t-Tau, (**E**) pTau181, and (**F**) Aβ42/pTau181. Shaded areas represent 95% CIs.

**Figure 3.**
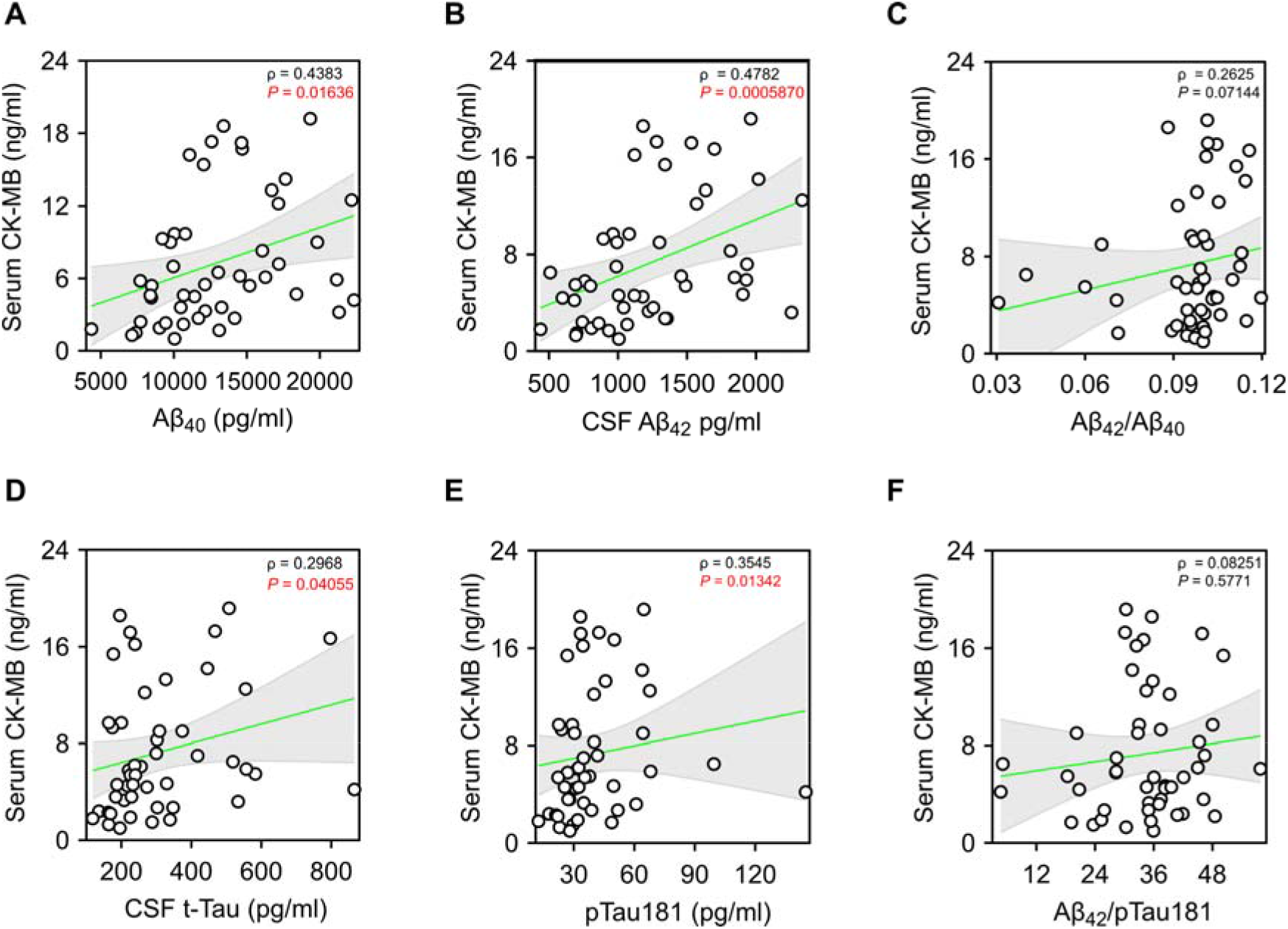
CSF AD Biomarkers Correlations with CK-MB. Correlation analyses were performed using nonparametric Spearman correlations (ρ). Curves were drawn by a linear regression model with an interaction term for serumCK-MB by CSF levels of (**A**) Aβ_40_, (**B**) Aβ_42_, (**C**) Aβ_42_/Aβ_40_ratio, (**D**) t-Tau, (**E**) pTau181, and (**F**) Aβ42/pTau181. Shaded areas represent 95% CIs.

### Correlations with pNfH and Cognitive Assessments

We next examined the relationship between canonical CSF AD biomarkers, markers of axonal degeneration, and cognitive performance in ALS. Using CSF phosphorylated neurofilament heavy chain (pNfH, n = 52) as an indicator of neuroaxonal injury (Figure 4 A-F) and the ECAS (n = 33), according to availability, as the principal measure of cognitive function (Figure 5 A-F). CSF pNfH concentrations showed significant negative correlations with both Aβ_42_ (ρ = –0.32, P = 0.021; Figure 4B) and the Aβ_42_/Aβ_40_ ratio (ρ = –0.29. P = 0.038;; Figure 4C). In a subset of participants with available ECAS data (n = 33), the Aβ_42_/Aβ_40_ ratio correlated positively with the non–ALS-specific memory sub-score (ρ = 0.35, P = 0.048; Figure 5C), whereas no significant associations were observed for other cognitive domains (Supplementary Table 2). Collectively, these findings suggest that higher CSF Aβ_42_ levels may be linked to lower axonal degeneration and relatively preserved memory performance, potentially indicating a more favorable functional and cognitive phenotype within the ALS collective.

**Figure 4.**
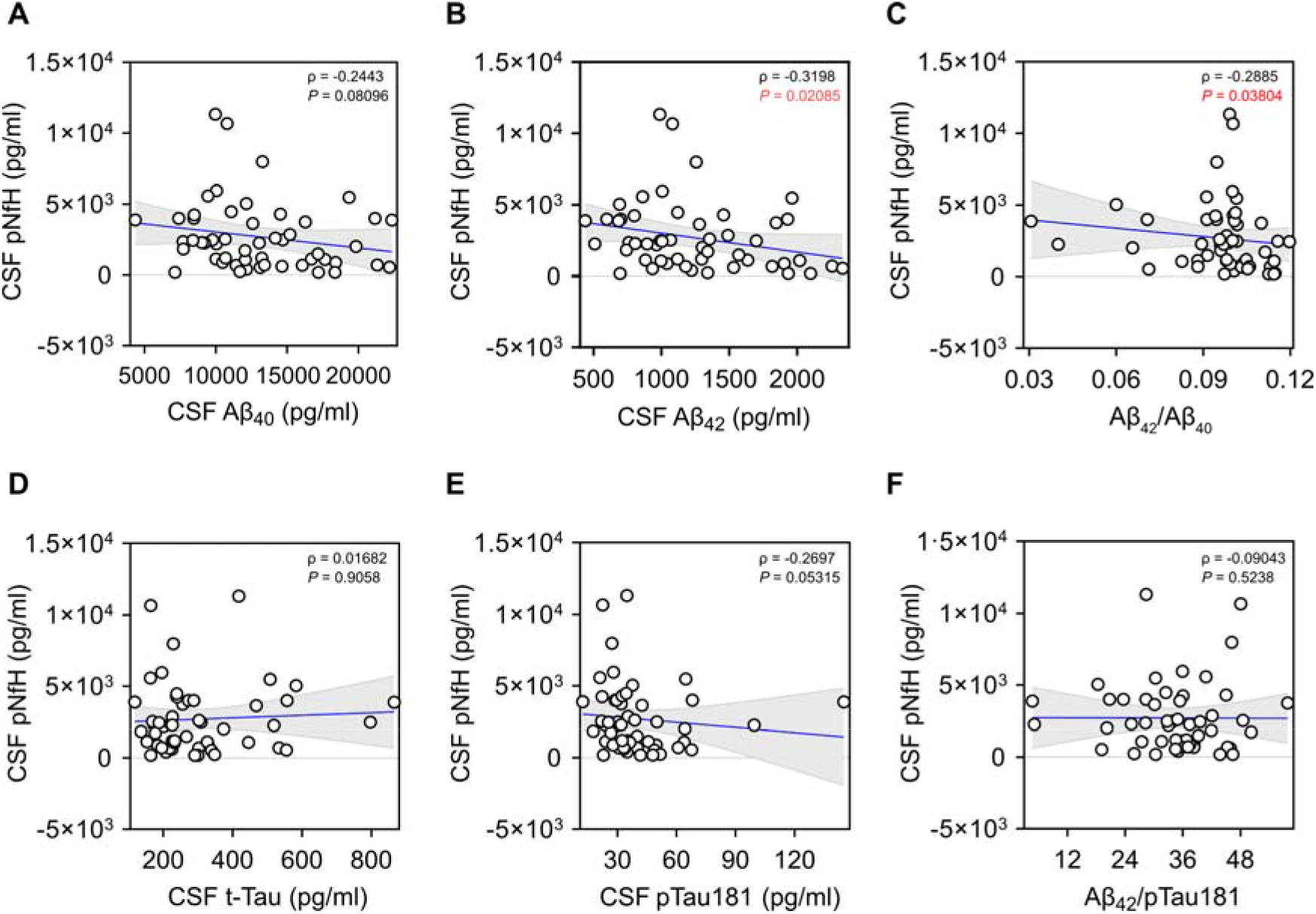
CSF AD Biomarkers Correlations with pNfH. Correlation analyses were performed using nonparametric Spearman correlations (ρ). Curves were drawn by a linear regression model with an interaction term for CSF pNfH by CSF levels of (**A**) Aβ_40_, (**B**) Aβ_42_, (**C**) Aβ_42_/Aβ_40_ratio, (**D**) t-Tau, (**E**) pTau181, and (**F**) Aβ42/pTau181. Shaded areas represent 95% CIs.

**Figure 5.**
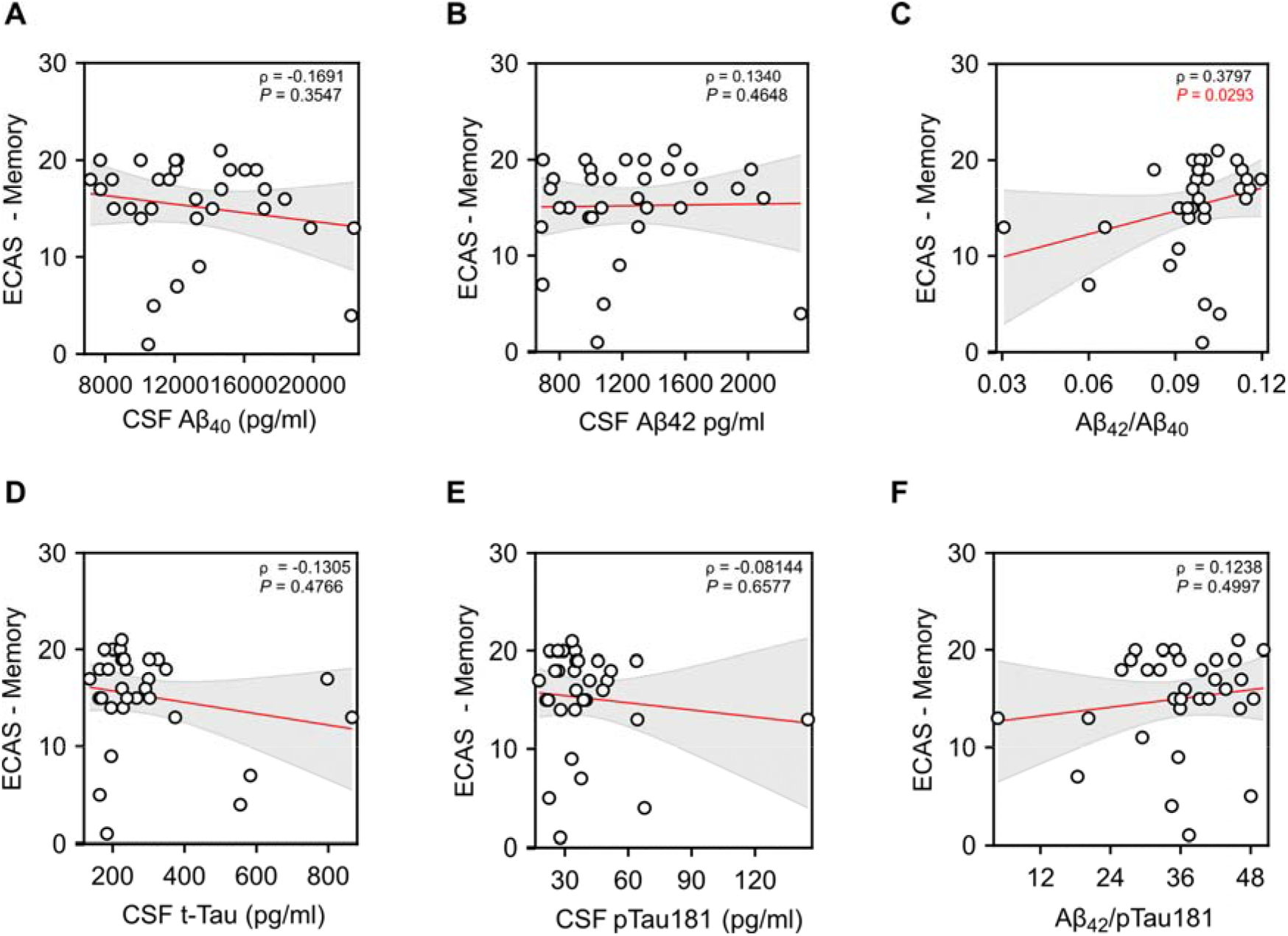
CSF AD Biomarkers Correlations with ECAS Memory score. Correlation analyses were performed using nonparametric Spearman correlations (ρ). Curves were drawn by a linear regression model with an interaction term for ECAS Memory score by CSF levels of (**A**) Aβ_40_, (**B**) Aβ_42_, (**C**) Aβ_42_/Aβ_40_ ratio, (**D**) t-Tau, (**E**) pTau_181_, and (**F**) Aβ42/pTau_181_. Shaded areas represent 95% CIs.

### Longitudinal CSF AD-related biomarker in Tofersen-Treated Patients

Having identified positive correlations between CSF AD biomarkers and functional and memory-related performance in ALS, we next conducted an exploratory, descriptive longitudinal analysis of these biomarkers in three patients with *SOD1*-associated ALS receiving the *SOD1* antisense oligonucleotide tofersen at our department. All three patients had initiated treatment before analysis; therefore, pretreatment biomarker measurements were unavailable. Biomarker measurements were obtained at intrathecal dosing visits, approximately every 30 days, over up to 12 months. For longitudinal visualization, each biomarker value was natural-logarithmically transformed; no additional normalization to baseline or to group mean was performed. The total number of measurements per patient is shown in Figure 6A. The log-transformed longitudinal trajectories showed a consistent downward trend in CSF Aβ_40_, Aβ_42_, t-Tau, and pTau181 (Figure 6A), with initial-to-final measurements showing a concordant pattern (Figure 6B–F). By contrast, CSF pNfH and plasma NfL levels appeared relatively stable (Figure 6G–H).

**Figure 6.**
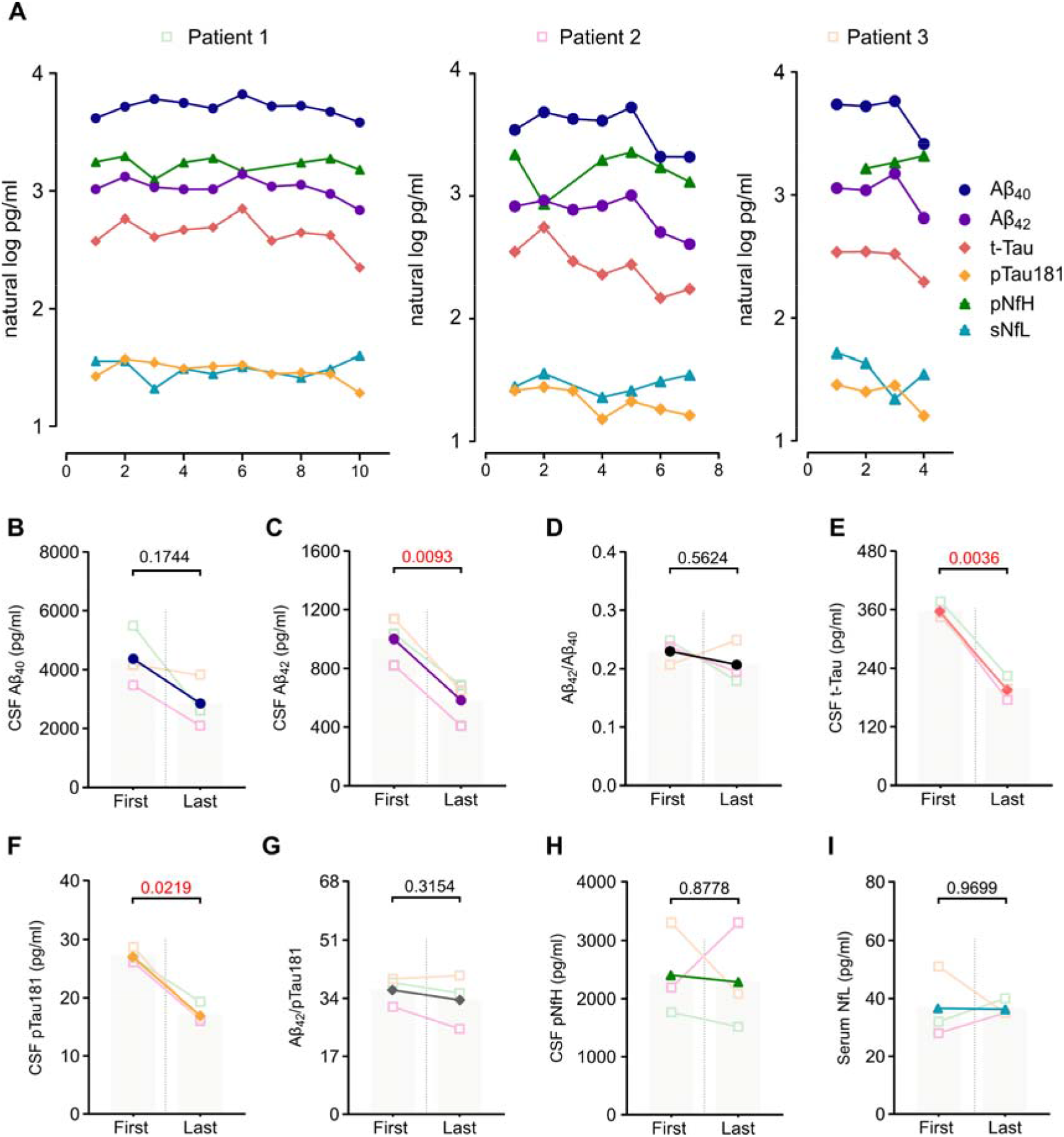
Longitudinal CSF AD-related biomarker in Tofersen-Treated Patients. (**A**) Longitudinal trajectories of natural log-transformed, normalized levels of AD-related biomarkers—CSF pNfH and plasma NfL—in three SOD1 mutation carriers treated with tofersen. Each dot represents an individual measurement taken on the day of intrathecal tofersen administration. Paired comparisons between the first and last measurements of mean CSF concentrations are shown fo (**B**) Aβ_40_, (**C**) Aβ_42_, (**D**) Aβ_42_/Aβ_40_ratio, (**E**) t-Tau, (**F**) pTau181, (**G**) pNfH and serum values of (**H**) NfL. Individual patient changes are represented by semi-transparent connecting lines. Reported *P* values are from paired *t*-tests.

These exploratory observations provide a preliminary longitudinal description of biomarker profiles during tofersen treatment. Although the absence of pretreatment measurements and the small sample size preclude attribution of these changes to treatment, the patterns may help inform future studies of disease-related biomarker trajectories and potential treatment-associated changes.

## Discussion

The identification of robust diagnostic and prognostic biomarkers in ALS remains an important unmet need. In this study, we evaluated the relevance of AD-associated CSF biomarkers in ALS by examining their diagnostic performance, associations with disease-related measures in a real-world cohort, and longitudinal profiles during tofersen treatment in three patients with SOD1-associated ALS. In this retrospective comparative study, CSF concentrations of Aβ_40_, Aβ_42_, t-Tau and pTau181 in patients with ALS were broadly comparable to those in individuals with non-neurodegenerative disorders, whereas the AD cohort showed the expected differences. These findings indicate that, within this cohort and assay framework, canonical AD biomarkers in CSF did not provide disease-specific diagnostic discrimination for ALS. Importantly, we identified biologically plausible associations with clinical severity, neurofilament concentrations, markers of muscle injury, and mnemonic performance. In the three patients with *SOD1*-mutant ALS receiving tofersen, the biomarkers showed directional changes over time, suggesting a potentially treatment-responsive effect.

The broadly similar CSF Aβ and Tau concentrations in the ALS and control cohorts are consistent with reports of minimal or absent AD biomarker signatures in sporadic and familial ALS^25.26^. Although amyloid and Tau pathologies may co-occur in ALS, particularly in older individuals[25] the CSF findings in this study cannot establish their presence or extent at the neuropathological level. Overall, our results suggest that canonical amyloid- and Tau-related CSF profiles were not prominent biochemical features of ALS in this cohort. At the same time, the observed associations between Aβ species, neurofilament concentrations, and clinical measures reveal potentially informative relationships that merit further investigation and may help refine biomarker-based phenotyping in ALS.

Higher Aβ_40_and Aβ_42_ concentrations were positively associated with better functional performance, as measured by the ALSFRS-R. This observation is compatible with the possibility that higher Aβ concentrations reflect preserved neuronal or synaptic integrity, a hypothesis also raised by experimental work in human neurons[26]. Because this association was cross-sectional, it cannot distinguish among potential explanations, including differences in disease stage, age, BMI or CSF protein concentration, as well as variation in Aβ production or clearance. The association between Aβ peptides and CK-MB may similarly reflect shared disease burden or systemic stress[27]. However, CK-MB is not specific to ALS, and non-neurological influences cannot be excluded. Amyloid Precursor Protein (APP) processing represents a particularly compelling avenue for future investigation, given evidence that APP and its cleavage products participate in axonal regeneration and neuromuscular maintenance^30.31^. A recent study reported elevated serum pTau_181_ and pTau_217_ concentrations in ALS, comparable to those observed in AD, and suggested a potential muscular origin[30, 31]. In this context, the positive association between CSF pTau_181_ and serum CK-MB in our cohort may reflect parallel responses to disease-related stress in lower motor neurons and muscle, providing a testable hypothesis for future studies.

The inverse correlation between Aβ_42_ and pNfH further highlights a potentially meaningful relationship between these biomarkers in our cohort. Neurofilament elevation is an established indicator of axonal injury and motor neuron loss in ALS and other neurodegenerative diseases [32]. Although the cross-sectional association does not establish the underlying process or its direction, it is consistent with a link between Aβ_42_-related biology and axonal integrity. Exploratory analyses also identified a modest association between higher CSF Aβ_42_/Aβ_40_ ratios and better ECAS memory performance. This domain-specific finding, observed in a subset of patients, provides an encouraging signal that warrants replication in larger cohorts. Previous fluid biomarkers, imaging, and neuropathological studies of ALS–FTD have primarily linked cortical degeneration and frontotemporal atrophy to Tau and TDP-43 pathology[33, 34] rather than to Aβ species. Our findings add to the growing interest in whether synaptic and AD–related fluid biomarkers, when interpreted alongside cognitive measures, may contribute to phenotypic and prognostic stratification in ALS. The positive association is consistent with a recent report in which lower Aβ_42_/Aβ_40_ ratios were associated with poorer memory performance in ALS[24]. It is further supported by observations in cognitively impaired ALS and ALS-FTD, where significantly lower CSF Aβ_42_/Aβ_40_ and Aβ_42_/pTau181 ratios were found relative to healthy controls[34]. In line with this possibility, a lower CSF Aβ_42_/Aβ_40_ratio has also been associated with shorter survival in ALS, indicating that amyloid-related measures may reflect aspects of clinical heterogeneity beyond motor decline[35]. Intriguingly, we did not observe significant associations between CSF T-tau or pTau181 and ECAS performance. This divergence may reflect several factors, including inter-assay variability, differences in pre-analytical handling, and the relatively restricted cognitive spectrum of our cohort, which did not include patients with overt ALS-FTD.

In three *SOD1*-mutant ALS patients treated with tofersen, longitudinal CSF measurements showed directional decreases in Aβ_40_, Aβ_42_, t-Tau and pTau_181_ without apparent changes in CSF pNfH or serum NfL. Previous clinical studies have reported sustained reductions in neurofilament levels following SOD1 silencing with tofersen[36]. Differences between those findings and our observations may reflect the small sample, sampling timing, assay variability, regression to the mean or instability in individual longitudinal trajectories. Nevertheless, the parallel directional changes in Aβ and Tau biomarkers raise the intriguing possibility that SOD1 lowering may influence non-canonical pathways, including APP processing, Tau phosphorylation or broader proteostatic mechanisms. Similar decreases in Aβ_40_ and Aβ_42_for up to six months after tofersen initiation have been reported using NULISA™-based targeted proteomics and LUMIPULSE™ immunoassays[16]. Our single-center real-world observations, obtained using commercially available assays, are directionally consistent with these reports and provide independent support for the possibility that disease-modifying treatment may alter the CSF biomarker milieu beyond its primary molecular targets. Together, these observations generate a clear rationale for prospective studies of biomarker dynamics during SOD1-targeted therapy.

Several limitations should be considered. The retrospective design, small number of patients in the treatment analysis and clinical heterogeneity limit the strength of mechanistic conclusions. The control cohort was heterogeneous and included individuals with non-neurodegenerative conditions, such as depression and normal-pressure hydrocephalus, which may affect comparability. The AD cohort was clinically selected and defined from the clinic rather than recruited as a population-based reference group; this should be considered when interpreting between-group differences. Correlations were not adjusted for age or disease duration, and p-values were not adjusted for multiple testing, consistent with the exploratory nature of the analyses. The absence of complementary neuroimaging or post-mortem data also limits anatomical and neuropathological interpretation of the CSF findings. Despite these limitations, assessing canonical AD biomarkers alongside ALS-related measures, including pNfH, ALSFRS-R and ECAS, provides a multidimensional view of biomarker relationships in this real-world cohort. Prospective longitudinal studies in larger, well-characterized cohorts—with comparator groups, standardized pre-analytical procedures and sensitive single- and multi-analyte assays—are needed to assess the reproducibility and clinical relevance of these findings. Such studies could determine whether changes in Aβ and Tau track with neurofilament dynamics, cognition or treatment exposure, and may ultimately support the development of more comprehensive biomarker panels for ALS.

## Acknowledgements

We thank the current and former directors of the Departments of Neurodegenerative Diseases, Cognitive Disorders, Gerontopsychiatry, Neuromuscular and Neuroimmunological Diseases at the University Hospital Bonn — Prof. Michael T. Heneka, Prof. Anja Schneider, Prof. Ulrich Wüllner, Prof. Cornelia Kornblum and Prof. Anne-Kathrin Pröbstel— for their support of this work.

## Author contributions

S.C-G., and P.W. conceived the study. F.E. und S.C-G. analysed data, interpreted results and performed statistical analyses. S.B., R.F., P.W., C.W. and S.C-G. acquired data and contributed to clinical expertise, F.E., S.C-G., S.B., C.W. critically revised the manuscript. S.C-G. and F.E. drafted the manuscript. All authors read and approved the manuscript.

## Data availability

All details regarding the presented datasets are available from the corresponding authors upon request.

## Supplementary material

**Supplementary Table 1.**
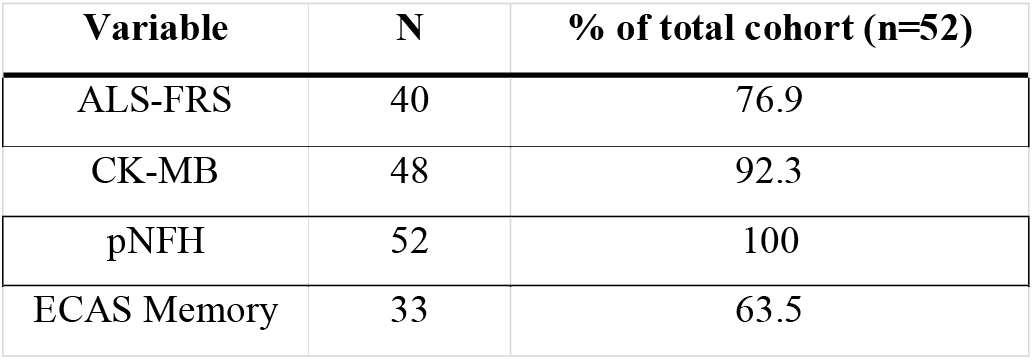
Date Availability.

| **Variable** | **N** | **% of total cohort (n=52)** |
|---|---|---|
| ALS-FRS | 40 | 76.9 |
| CK-MB | 48 | 92.3 |
| pNFH | 52 | 100 |
| ECAS Memory | 33 | 63.5 |

**Supplementary Table 2.** Correlations CSF AD Biomarkers and ECAS.

|  | **Global** |  | **ALS-S** |  | **ALS-NS** |  | **Language** |  | **Speech** |  | **Executive** |  | **Memory** |  | Spatial |  |
|---|---|---|---|---|---|---|---|---|---|---|---|---|---|---|---|---|
| **Marker** | **ρ** | ***P*-value** | **ρ** | ***P*-value** | **ρ** | ***P*-value** | **ρ** | ***P*-value** | **ρ** | ***P*-value** | **ρ** | ***P*-value** | **ρ** | ***P*-value** | **ρ** | ***P*-value** |
| **Aβ<sub>40</sub>** | 0.08 | 0.6803 | 0.09 | 0.6148 | 0.01 | 0.9689 | 0.09 | 0.6025 | 0.07 | 0.7175 | -0.04 | 0.8141 | -0.17 | 0.3547 | 0.19 | 0.2714 |
| **Aβ<sub>42</sub>** | 0.24 | 0.172 | 0.21 | 0.2608 | 0.25 | 0.159 | 0.03 | 0.8501 | 0.09 | 0.6407 | 0.14 | 0.454 | 0.13 | 0.4648 | 0.15 | 0.3953 |
| **t-Tau** | -0.07 | 0.7218 | -0.08 | 0.6682 | -0.06 | 0.7626 | 0.06 | 0.7147 | -0.08 | 0.6569 | -0.2 | 0.2574 | -0.13 | 0.4766 | -0.04 | 0.8301 |
| **pTau181** | 0.02 | 0.9333 | 0.02 | 0.9296 | 0.02 | 0.9158 | 0.09 | 0.6071 | 0.05 | 0.7892 | -0.135 | 0.4617 | -0.085 | 0.6577 | 0.04 | 0.8127 |
| **Aβ<sub>42</sub>/Aβ<sub>40</sub>** | 0.34 | *0.0507* | 0.28 | 0.1187 | 0.33 | 0.0544 | 0.11 | 0.5179 | 0.2 | 0.2635 | 0.22 | 0.2137 | 0.38 | *\*0.0293* | 0.14 | 0.401 |
| **Aβ<sub>42</sub>/pTau181** | 0.26 | 0.1528 | 0.29 | 0.1029 | 0.2 | 0.2587 | 0.05 | 0.7702 | 0.22 | 0.2289 | 0.27 | 0.2057 | 0.12 | 0.4997 | 0.31 | 0.0778 |

## References

1. Vidovic M, Müschen LH, Brakemeier S, Machetanz G, Naumann M, Castro-Gomez S. Current State and Future Directions in the Diagnosis of Amyotrophic Lateral Sclerosis. Cells. 2023;12:736. 10.3390/cells12050736.

2. Ravits J, Ferrey D, Gundogdu B, Qayoumi W, Zale C. Amyotrophic Lateral Sclerosis: A Review. JAMA. 2026;335:1970. 10.1001/jama.2026.6385.

3. Masrori P, Van Damme P. Amyotrophic lateral sclerosis: a clinical review. Eur J Neurol. 2020;27:1918–29. 10.1111/ene.14393.

4. de Carvalho M, Dengler R, Eisen A, England JD, Kaji R, Kimura J, et al. Electrodiagnostic criteria for diagnosis of ALS. Clin Neurophysiol. 2008;119:497–503. 10.1016/j.clinph.2007.09.143.

5. Shefner JM, Al-Chalabi A, Baker MR, Cui L-Y, De Carvalho M, Eisen A, et al. A proposal for new diagnostic criteria for ALS. Clinical Neurophysiology. 2020;131:1975–8. 10.1016/j.clinph.2020.04.005.

6. Cedarbaum JM, Stambler N, Malta E, Fuller C, Hilt D, Thurmond B, et al. The ALSFRS-R: a revised ALS functional rating scale that incorporates assessments of respiratory function. BDNF ALS Study Group (Phase III). J Neurol Sci. 1999;169:13–21. 10.1016/s0022-510x(99)00210-5.

7. Pinto-Grau M, Burke T, Lonergan K, McHugh C, Mays I, Madden C, et al. Screening for cognitive dysfunction in ALS: validation of the Edinburgh Cognitive and Behavioural ALS Screen (ECAS) using age and education adjusted normative data. Amyotroph Lateral Scler Frontotemporal Degener. 2017;18:99–106. 10.1080/21678421.2016.1249887.

8. Gaiani A, Martinelli I, Bello L, Querin G, Puthenparampil M, Ruggero S, et al. Diagnostic and Prognostic Biomarkers in Amyotrophic Lateral Sclerosis: Neurofilament Light Chain Levels in Definite Subtypes of Disease. JAMA Neurol. 2017;74:525. 10.1001/jamaneurol.2016.5398.

9. Verde F, Steinacker P, Weishaupt JH, Kassubek J, Oeckl P, Halbgebauer S, et al. Neurofilament light chain in serum for the diagnosis of amyotrophic lateral sclerosis. J Neurol Neurosurg Psychiatry. 2019;90:157–64. 10.1136/jnnp-2018-318704.

10. De Schaepdryver M, Jeromin A, Gille B, Claeys KG, Herbst V, Brix B, et al. Comparison of elevated phosphorylated neurofilament heavy chains in serum and cerebrospinal fluid of patients with amyotrophic lateral sclerosis. J Neurol Neurosurg Psychiatry. 2018;89:367–73. 10.1136/jnnp-2017-316605.

11. Verde F, Zaina G, Bodio C, Borghi MO, Soranna D, Peverelli S, et al. Cerebrospinal fluid phosphorylated neurofilament heavy chain and chitotriosidase in primary lateral sclerosis. J Neurol Neurosurg Psychiatry. 2021;92:221–3. 10.1136/jnnp-2020-324059.

12. Khalil M, Teunissen CE, Lehmann S, Otto M, Piehl F, Ziemssen T, et al. Neurofilaments as biomarkers in neurological disorders - towards clinical application. Nat Rev Neurol. 2024;20:269–87. 10.1038/s41582-024-00955-x.

13. Hamilton RL, Bowser R. Alzheimer disease pathology in amyotrophic lateral sclerosis. Acta Neuropathol. 2004;107:515–22. 10.1007/s00401-004-0843-1.

14. Matías-Guiu JA, Pytel V, Cabrera-Martín MN, Galán L, Valles-Salgado M, Guerrero A, et al. Amyloid- and FDG-PET imaging in amyotrophic lateral sclerosis. Eur J Nucl Med Mol Imaging. 2016;43:2050–60. 10.1007/s00259-016-3434-1.

15. Chen Y, Sun S, Bai Z, Gao N, Yu W, Sun X, et al. CSF Aβ, Tau, Axonal, Synaptic, Glial, Neural, and Inflammatory Biomarkers in Patients With Sporadic Amyotrophic Lateral Sclerosis. Neurology. 2025;105:e213914. 10.1212/WNL.0000000000213914.

16. Steffke C, Baskar K, Bachhuber F, Wiesenfarth M, Dorst J, Schuster J, et al. Targeted Proteomics upon Treatment with Tofersen Identifies Novel Response Markers for Superoxide Dismutase 1-Linked Amyotrophic Lateral Sclerosis. Ann Neurol. 2025;98:1318–34. 10.1002/ana.70025.

17. Weishaupt JH, Körtvélyessy P, Schumann P, Valkadinov I, Weyen U, Hesebeck-Brinckmann J, et al. Tofersen decreases neurofilament levels supporting the pathogenesis of the SOD1 p.D91A variant in amyotrophic lateral sclerosis patients. Commun Med (Lond). 2024;4:150. 10.1038/s43856-024-00573-0.

18. Ludolph A, Drory V, Hardiman O, Nakano I, Ravits J, Robberecht W, et al. A revision of the El Escorial criteria - 2015. Amyotrophic Lateral Sclerosis and Frontotemporal Degeneration. 2015;16:291–2. 10.3109/21678421.2015.1049183.

19. Abdulla S, Vielhaber S, Körner S, Machts J, Heinze H-J, Dengler R, et al. Validation of the German version of the extended ALS functional rating scale as a patient-reported outcome measure. J Neurol. 2013;260:2242–55. 10.1007/s00415-013-6955-6.

20. Abrahams S, Newton J, Niven E, Foley J, Bak TH. Screening for cognition and behaviour changes in ALS. Amyotroph Lateral Scler Frontotemporal Degener. 2014;15:9–14. 10.3109/21678421.2013.805784.

21. Halbgebauer S, Steinacker P, Verde F, Weishaupt J, Oeckl P, Von Arnim C, et al. Comparison of CSF and serum neurofilament light and heavy chain as differential diagnostic biomarkers for ALS. J Neurol Neurosurg Psychiatry. 2022;93:68–74. 10.1136/jnnp-2021-327129.

22. Gobom J, Parnetti L, Rosa-Neto P, Vyhnalek M, Gauthier S, Cataldi S, et al. Validation of the LUMIPULSE automated immunoassay for the measurement of core AD biomarkers in cerebrospinal fluid. Clin Chem Lab Med. 2022;60:207–19. 10.1515/cclm-2021-0651.

23. Agah E, Mojtabavi H, Behkar A, Heidari A, Ajdari A, Shaka Z, et al. CSF and blood levels of Neurofilaments, T-Tau, P-Tau, and Abeta-42 in amyotrophic lateral sclerosis: a systematic review and meta-analysis. J Transl Med. 2024;22:953. 10.1186/s12967-024-05767-7.

24. Maranzano A, Verde F, Dubini A, Torre S, Colombo E, Doretti A, et al. Association of APOE genotype and cerebrospinal fluid Aβ and tau biomarkers with cognitive and motor phenotype in amyotrophic lateral sclerosis. Eur J Neurol. 2024;31:e16374. 10.1111/ene.16374.

25. Moriyoshi H, Akagi A, Riku Y, Sone J, Miyahara H, Yoshida M, et al. Contribution of comorbid pathologies to amyotrophic lateral sclerosis with cognitive or behavioral abnormalities. BMC Neurol. 2025;26:19. 10.1186/s12883-025-04556-z.

26. Siddu A, Natale S, Wong CH, Shaye H, Südhof TC. Aggregation shifts amyloid-β peptides from synaptogenic to synaptotoxic. J Clin Invest. 2025;135:e193407. 10.1172/JCI193407.

27. Kittipeerapat N, Fabian R, Bernsen S, Weydt P, Castro-Gomez S. Creatine Kinase MB Isoenzyme Is a Complementary Biomarker in Amyotrophic Lateral Sclerosis. Int J Mol Sci. 2023;24:11682. 10.3390/ijms241411682.

28. Vohra BPS, Sasaki Y, Miller BR, Chang J, DiAntonio A, Milbrandt J. Amyloid precursor protein cleavage-dependent and -independent axonal degeneration programs share a common nicotinamide mononucleotide adenylyltransferase 1-sensitive pathway. J Neurosci. 2010;30:13729–38. 10.1523/JNEUROSCI.2939-10.2010.

29. Wang P, Yang G, Mosier DR, Chang P, Zaidi T, Gong Y-D, et al. Defective neuromuscular synapses in mice lacking amyloid precursor protein (APP) and APP-Like protein 2. J Neurosci. 2005;25:1219–25. 10.1523/JNEUROSCI.4660-04.2005.

30. Abu-Rumeileh S, Scholle L, Mensch A, Großkopf H, Ratti A, Kölsch A, et al. Phosphorylated tau 181 and 217 are elevated in serum and muscle of patients with amyotrophic lateral sclerosis. Nat Commun. 2025;16:2019. 10.1038/s41467-025-57144-7.

31. Abu-Rumeileh S, Großkopf H, Guthknecht A, Mensch A, Osterloh D, Ratti A, et al. Tau isoform mapping across tissues and biofluids reveals a signature distinguishing Alzheimer’s disease from amyotrophic lateral sclerosis. Nat Commun. 2026;17:10745. 10.1038/s41467-026-78390-3.

32. Poesen K, Van Damme P. Diagnostic and Prognostic Performance of Neurofilaments in ALS. Front Neurol. 2018;9:1167. 10.3389/fneur.2018.01167.

33. Chatterjee M, Özdemir S, Fritz C, Möbius W, Kleineidam L, Mandelkow E, et al. Plasma extracellular vesicle tau and TDP-43 as diagnostic biomarkers in FTD and ALS. Nat Med. 2024;30:1771–83. 10.1038/s41591-024-02937-4.

34. Kasper E, Lehto A, Jürs A, Nordmann N, Peters O, Hellmann J, et al. Alzheimer’s Disease Co Pathology and Cognitive Impairment in Amyotrophic Lateral Sclerosis. Annals of Neurology. 2026;100:123–38. 10.1002/ana.78227.

35. Colletti T, Agnello L, Spataro R, Guccione L, Notaro A, Lo Sasso B, et al. Prognostic Role of CSF β-amyloid 1-42/1-40 Ratio in Patients Affected by Amyotrophic Lateral Sclerosis. Brain Sci. 2021;11:302. 10.3390/brainsci11030302.

36. Miller T, Cudkowicz M, Shaw PJ, Andersen PM, Atassi N, Bucelli RC, et al. Phase 1–2 Trial of Antisense Oligonucleotide Tofersen for *SOD1* ALS. N Engl J Med. 2020;383:109–19. 10.1056/NEJMoa2003715.

